# Diet, pace of biological aging, and risk of dementia in the Framingham Heart Study

**DOI:** 10.1101/2023.05.24.23290474

**Authors:** Aline Thomas, Calen P Ryan, Avshalom Caspi, Terrie E. Moffitt, Karen Sugden, Jiayi Zhou, Daniel W. Belsky, Yian Gu

## Abstract

**INTRODUCTION:** We tested the hypothesis that healthy diet protects against dementia because it slows the pace of biological aging.

**METHODS:** We analyzed Framingham Offspring Cohort data (≥60y). We measured healthy diet using the Dietary Guideline for Americans (DGA, 3 visits 1991-2008), pace of aging using the DunedinPACE epigenetic clock (2005-2008), and incident dementia and mortality using records (compiled 2005-2018).

**RESULTS:** Of n=1,525 included participants (mean age 69.7, 54% female), n=129 developed dementia and n=432 died over follow-up. Greater DGA adherence was associated with slower DunedinPACE and reduced risks for dementia and mortality. Slower DunedinPACE was associated with reduced risks for dementia and mortality. Slower DunedinPACE accounted for 15% of the DGA association with dementia and 39% of the DGA association with mortality.

**DISCUSSION:** Findings suggest that slower pace of aging mediates part of the relationship of healthy diet with reduced dementia risk. Monitoring pace of aging may inform dementia prevention.

## 1. Introduction

People who eat healthier diets are less likely to develop dementia.^1–3^ However, the biological mechanism of this protection is not well understood.^4^ By far the greatest risk factor for dementia is aging. Biological processes linked with healthy diet and reduced dementia risk include reversal of several so-called hallmarks of aging, including metabolic regulation and reduced inflammation and oxidative stress, among others.^5–8^ These observations, together with data from animal model and human observational studies linking healthy diet with health-span and lifespan,^9^ suggest the hypothesis that one mechanism through which healthy diet reduces dementia risk is a slowing of processes of biological aging.

Biological aging is the progressive decline in system integrity that occurs with advancing age.^10^ It arises from the accumulation of molecular changes that undermine the functioning and resilience capacity of tissues and organs, ultimately causing disease and death.^11, 12^ Recent advances in aging research have produced new measurements of this process.^13^ The best-validated among these new measures of aging are algorithms that combine information from dozens to hundreds of chemical tags, called methylation marks, on the DNA sequence of white blood cells to estimate the pace and progress of biological aging.^14^ These algorithms are known as epigenetic clocks. Several epigenetic clocks, in particular the PhenoAge, GrimAge, and DunedinPACE clocks, have accumulated substantial evidence as biomarkers predictive of health-span and lifespan.^15–17^ These clocks also tend to indicate younger biological age and slower Pace of Aging in people who eat healthier diets.^18–21^ In our study, we focused on DunedinPACE because evidence for associations with cognitive and brain aging outcomes were more consistent for DunedinPACE than for the PhenoAge and GrimAge clocks, including in the Framingham Offspring Cohort studied here.^17, 22–25^

We analyzed data on diet, biological aging, and incidence of dementia collected over three decades of follow-up in the Framingham Heart Study Offspring Cohort. We assessed long-term healthy diet by the Dietary Guidelines Adherence Index (DGAI). We measured pace of biological aging using the DunedinPACE clock. Dementia-free survival was determined from the Framingham Study’s records. We tested if participants with a healthier diet had slower Pace of Aging and improved dementia-free survival and if a slower Pace of Aging mediated the diet-dementia association. We also explored the specificity of this pathway for brain aging, as compared to overall aging, by contrasting the proportion mediated for dementia to the proportion mediated for all-cause mortality.

## 2. Methods

### 2.1. Study population

The Framingham Heart Study is an ongoing population-based cohort following three generations of families recruited, starting 1948, within the town of Framingham, Massachusetts, USA.^26^ We analyzed data from the second generation of participants, the Offspring Cohort. Initiated in 1971, The Offspring Cohort was initiated in 1971 and participants have since been followed-up at nine examinations, approximately every 4-7 years.^27^ At each follow-up visit, data collection included physical examination, lifestyle-related questionnaires, blood sampling, and, starting in 1991, neurocognitive testing.^28^ The study protocol was approved by the institutional Review Board for Human Research at Boston University Medical Center, and all participants provided written inform consent. Data for the Framingham Offspring Study were obtained from dbGaP (phs000007.v33.p14).

### 2.2. DNA methylation

Whole-genome DNA methylation (DNAm) profiles were obtained from dbGaP (phs000724.v10.p14). For the Offspring Cohort, DNAm was measured from whole-blood buffy-coat samples collected at Visit 8 (2005-2008) using the Infinium HumanMethylation450 BeadChip (Illumina). Processing and normalization of DNAm data has been described previously.^29^ Briefly, data were normalized using the “dasen” method in the ‘wateRmelon’ R package^30^ and subjected to downstream QC. Samples with missing rate >1% at p<0.01, poor SNP matching to the 65 SNP control probe locations, and outliers by multidimensional scaling techniques were excluded. Probes with missing rate of >20% at p<0.01 were also excluded.

The DunedinPACE epigenetic clock was computed from the DNAm dataset using the code available on GitHub (https://github.com/danbelsky/DunedinPACE) following the procedure described in Belsky and al.^17^ Briefly, DunedinPACE was developed in the Dunedin Longitudinal Study from analysis of the Pace of Aging phenotype. The Dunedin Study follows a cohort of 1,037 births at the Queen Mary Hospital in Dunedin, New Zealand in 1972-73. The cohort has been followed up through members’ 45 birthdays with 95% retention among survivors.^31^ Pace of Aging is an integrative summary of rate-of-change estimates for 19 blood biomarker and organ-function-tests.^25, 32^ Rate-of-change estimates were derived from longitudinal analysis of measurements collected across 4 study visits at ages 26, 32, 38, and 45. DunedinPACE was computed from elastic-net regression of Pace of Aging on blood DNAm data collected from participants at age 45. The resulting algorithm included 173 cytosine-phosphate-guanine (CpG) sites. We computed DunedinPACE for Framingham participants using the publicly available R package and the DNA methylation dataset published by the Framingham Heart Study on NIH dbGaP (accession phs000724.v10.p14). DunedinPACE values are normed to the Dunedin Study birth cohort, in which a value of 1 corresponds to expected biological decline per calendar year over the age 26-45 follow-up interval. Values greater than 1 biological year per chronological year indicate a faster Pace of Aging. Values below 1 indicate a slower Pace of Aging. For analysis, the DunedinPACE values were standardized to mean = 0, standard deviation (SD) = 1, so that positive values indicate faster-than-average Pace of Aging within the Framingham Offspring cohort and negative values indicate slower-than-average Pace of Aging. Standardized DunedinPACE was analyzed as a continuous variable but categorized by tertiles for descriptive analyses.

### 2.3. Dietary assessment

The Offspring Cohort collected dietary information at each study visit using the validated 126-item Harvard semi-quantitative Food Frequency Questionnaire (FFQ), recording habitual food consumptions over the past year.^33, 34^ The frequency of consumption of each food items (i.e., from never or <1 per month to >6 per day) was converted into servings-per-week using common portion size. Nutrient intakes were estimated by multiplying the frequency of consumption of each food item by the nutrient content of the specified portions.

The 2010 Dietary Guideline Adherence Index (DGAI) assesses the conformity to the U.S. 2010 Dietary Guidelines for Americans (DGA).^35^ The detailed methodology for the development of the DGAI has been presented elsewhere.^36^ Briefly, the DGAI score is composed of two subscores: energy-specific food intake and healthy choice. The energy-specific food intake subscore measures adherence to energy-dependent DGA recommendations for intake and variety of 14 food components: fruit; dark green vegetables; orange/red vegetables; starchy vegetables; other vegetables; grains; dairy; meat/proteins/eggs; seafood; nuts/seeds/soy; legumes; sugar; variety in protein choices; and variety of fruits and vegetables. For this subscore, recommendations are defined for each individual depending on specific levels of energy requirements based on participant’s age, sex, body mass index, and physical activity estimates.^37^ The healthy choice subscore measures the conformity to DGA recommendations regarding consumptions of 11 quality food groups and nutrients. It includes consumption recommendation for 7 nutrients (total fat, saturated fat, *trans* fat, cholesterol, sodium, fiber, and alcohol) and the percentage of quality food consumed within 4 food groups (lean protein, low-fat dairy, whole grain, whole fruits). Conformity to recommendations for each subscore component was scored proportionally on a continuous scale of 0-1, with the specificity of applying a penalty for overconsumption of energy-dense foods (>50 kcal/serving) to prevent individuals from obtaining higher DGAI scores solely by consuming more food. The final DGAI score was obtained by summing component scores and standardizing the final score to a 0-100 points scale. A higher DGAI score indicates higher conformity to the 2010 DGA.

We computed participants’ DGAI scores from data collected at the 5^th^ (1991-95), 7^th^ (1998-2001) and 8^th^ (2005-2008) assessments (physical activity estimates for computation of energy-dependent requirements were not assessed at Visit 6). For analysis, DGAI scores were averaged across the three examination cycles to measure participants’ long-term adherence to healthy eating guidelines (66% of participants had completed FFQs at all three exams, and the rest had two FFQs). The DGAI score was analyzed as a continuous variable and categorized by tertiles for descriptive analyses.

### 2.4. Dementia diagnosis

Dementia diagnosis was made following a three-step procedure.^38–40^ First, participants were flagged for further examination if (1) they performed lower than the education-based cut-off scores for Mini Mental State Examination assessed at each visit; (2) subjective cognitive impairment was reported by the participant or a family member; (3) referred by a treating physician or by an ancillary investigator of the cohort; or (4) after review of outside medical records. Second, flagged participants underwent additional annual neurologic and neuropsychological examinations. Third, all cases of possible cognitive decline and dementia were reviewed by a committee to determine the presence of dementia based on criteria of the Diagnostic and Statistical Manual of Mental Disorders-IV.^41^ Date of onset was determined by the committee after reviewing the systematical tracking of cognitive status before and after dementia diagnosis. Incident cases were adjudicated until year 2018.

### 2.5. Mortality data

All-cause mortality data were obtained from continuous surveillance of medical and hospital records, death certificates, communication with personal physicians, and next-of-kin interviews. Three physicians reviewed and adjudicated the information until year 2018.

### 2.6. Covariates

Demographic, socioeconomic and lifestyle variables were collected at Visit 8, and included age, sex, educational level (less than high school; high school graduate; some college; college graduate), marital status (never married; married or cohabitating; separated, divorced, or widowed). Apolipoprotein E (*APOE*) LJ4 allele carrier status was considered dichotomously (carrying at least one LJ4 allele versus no LJ4 allele). Body Mass Index (BMI) was calculated as weight divided by height squared (in kg/m²; continuous). A physical activity index was computed as a composite score of time spend in five type of activities, assessed by a physical activity questionnaire, weighted by their intensity level (i.e., numbers of hours per day spend sleeping [weight factor = 1], in sedentary [weight factor = 1.1], light [weight factor = 1.5], moderate [weight factor = 2.4] or heavy [weight factor = 5.0] activities).^42^ Smoking status was defined as current if the individual had reported smoking during the year preceding baseline examination, as former if the individual had reported smoking at any previous examination visit, and never otherwise. Histories of cardiovascular disease and diabetes were self-reported at each visit. White blood cell composition was estimated from the DNAm data using the algorithms developed by Houseman et al. to estimate relative abundances of CD4 T cells, CD8 T cells, natural killer cells, B lymphocytes, monocytes, and granulocytes.^43^

### 2.7. Statistical analysis

Our analysis examined the potential mediating role of a slower pace of biological aging linking healthy diet with reduced risk of dementia. For comparative purposes, we conducted parallel analysis with all-cause mortality as the outcome.

### Analytic sample

For the present study, baseline was set at Visit 8, when DNAm to compute the DunedinPACE measure of Pace of Aging (mediator) was collected. Dietary data (exposure) were collected during the 16-year period prior to baseline (Visits 5 [1991-95], 7 [1998-2001], and 8 [2005-2008]). Follow-up for dementia and mortality (outcomes) were conducted from Visit 8 through 2018, the most recent date for which data were available. Our analysis sample included participants who were aged 60 or older and free of dementia at baseline Visit 8, who completed at least two FFQs at the Visits 5, 7 and/or 8, for whom DNAm data were available, and who were followed-up to ascertain dementia status.

The Offspring Cohort Visit 8 included 2,798 participants, of whom 2,479 (89%) had DNAm data passing quality control (**Figure 1**). Among them, 1,871 (75%) participants were over 60 years-old. We excluded participants not followed for dementia (n=2), with prevalent dementia at baseline (n=160), or with less than two valid FFQs (either because dietary intake data not available, or an abnormal estimated total energy intake [<600 or >3999 kcal for women; or <600 or >4199 kcal for men] and/or >13 missing items;^44^ n=184). The final sample for analysis included 1,525 participants.

**Figure 1.**
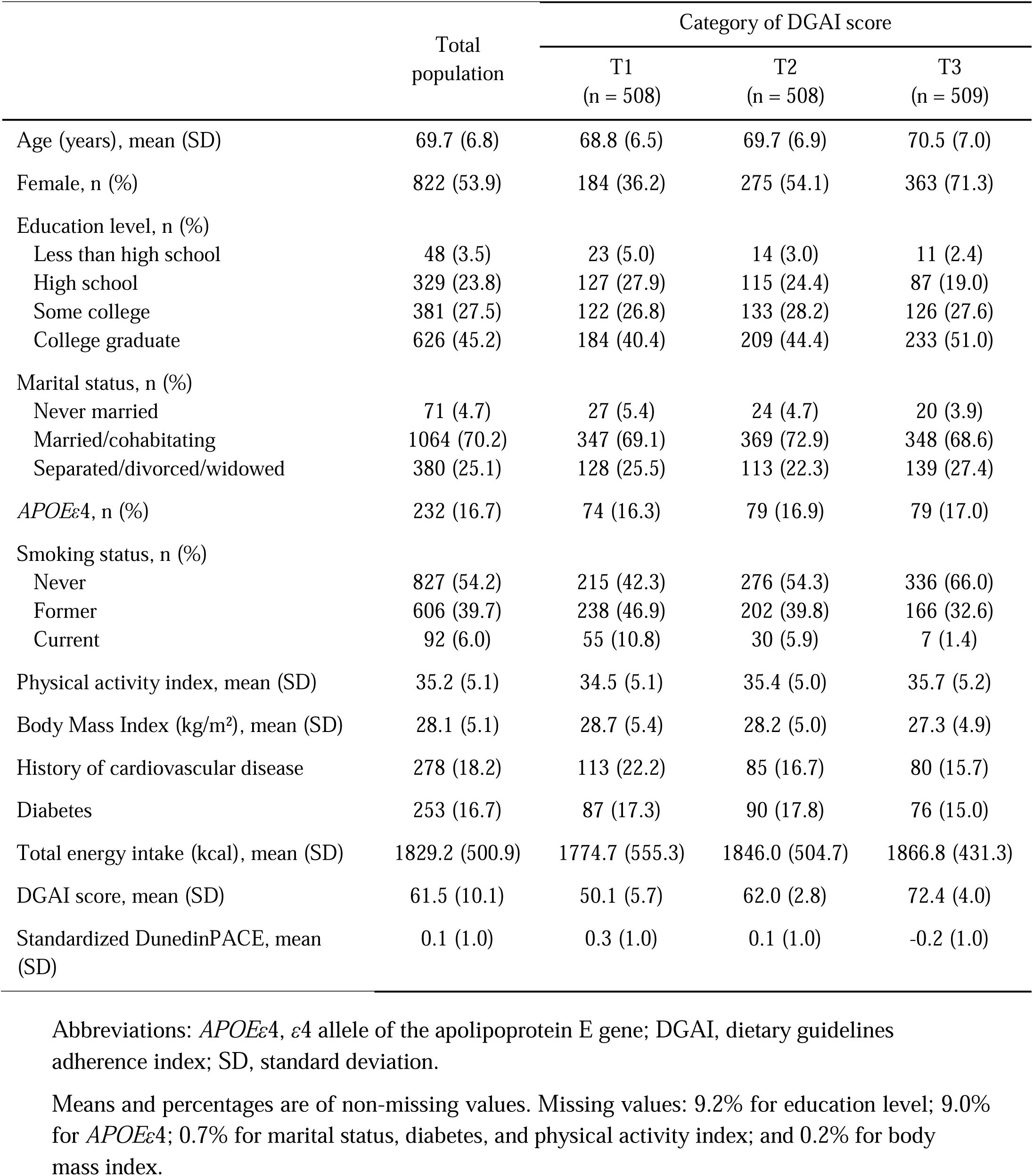
Flowchart of the study participants.

### Analysis of mediation by Pace of Aging

Mediation analysis was performed using the ‘CMAverse’ R package.^45^ First, the total effect of DGAI score (exposure) on dementia (outcome), i.e., coefficient {3_C_ and corresponding hazard ratio HR_C_, were estimated by a Cox proportional hazard model with time since baseline as time scale, adjusted for covariates. Second, the coefficient {3_a_ for the association of DGAI score with DunedinPACE (mediator) was estimated by adjusted linear regression. Third, the independent effects of the exposure and the mediator on dementia were estimated by a mutually adjusted Cox proportional hazard model and controlling for covariates, yielding the coefficient {3_b_ and resulting HR_b_ for the association of DunedinPACE with dementia, and coefficient {3_C1_ and HR_C1_ for the direct effect of DGAI score on dementia. Fourth, the indirect effect of DGAI on dementia via DunedinPACE was calculated as the exponential of {3_a_ x {3_b_, and the proportion of the diet-dementia association mediated by Pace of Aging was computed as ({3_a_ x {3_b_)⁄{3_C_. The 95% confident intervals (CI) for the indirect effect and the proportion mediated were estimated by 10,000 bootstrap samples in the R function procedure. The same mediation analysis was run with all-cause mortality as the outcome.

The same covariates were included in all models: age, age squared, sex, total energy intake, and the DNA methylation processing facility. Further adjustments controlling for socioeconomic factors (education level and marital status), for *APOE*LJ4 status, for other lifestyle factors (physical activity index, BMI, and smoking status), for history of medical conditions (cardiovascular disease and diabetes), or for blood cell composition to account for technical variation were evaluated by five models in sensitivity analysis.

### Supplementary analyses

We ran a series of supplementary analyses. First, we evaluated the modification effect of diet by sex, *APOE*LJ4 status, and smoking status (which was previously shown to be associated with DNA methylation profiles and to interact with diet in its association with biological aging).^19^ Second, we considered proximate and distant dietary habits by studying DGAI scores at each time point (i.e. at Visits 5, 7 and 8 separately). Third, we investigated adherence to a Mediterranean diet as an alternative healthy dietary pattern (see **Supplementary material** for computation details).^46^ Fourth, we explored two alternative DNA methylation measures of biological age: PhenoAge and GrimAge (see **Supplementary material** for computation details).^15, 16^

Statistical analyses were performed using R version 4.2.0 (R Foundation for Statistical Computing, Vienna, Austria). Analyses were performed on participants with all available covariates (none of which was missing for the primary model). The significance level was set at 0.05 for all tests. In Cox models, the log-linearity hypothesis was assessed and validated using restricted cubic splines,^47^ and the proportional-hazards assumption was investigated with Schoenfeld residuals.

## 3. Results

Among the 1,525 participants included, 54% were women and the mean age was 69.7 (±6.8) years (**Table 1**). The mean DGAI score was 61.5 (±10.1) points. Participants with higher adherence to dietary guidelines were older, had higher educational level, were more often married, were less likely to be smokers, practiced more physical activity, and had lower BMI.

**Table 1.**
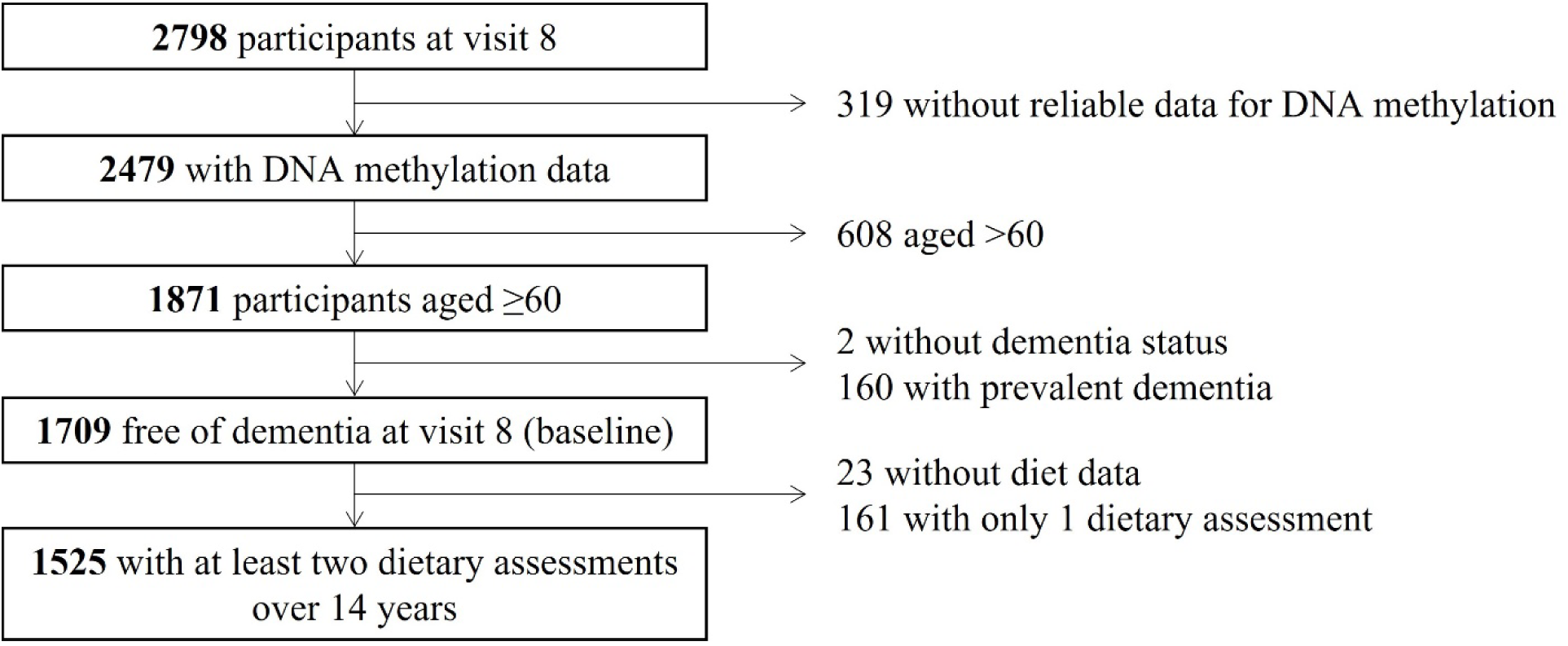
Characteristics of the study participants, The Framingham Offspring cohort, 1991-2018 (n = 1,525)

### 3.1. Diet, Pace of Aging, and risk of dementia

Over a median follow-up of 8.5 years (with maximum 13.4 years, and a total of 12,986 person-years), 129 participants (8.5%) were diagnosed with dementia. Participants with a faster Pace of Aging at baseline had an increased risk of developing dementia (HR_b_ [95% CI] = 1.34 [1.13; 1.60] for each 1-SD difference in DunedinPACE) (**Figure 2, panel C** and **Figure 3, panel A**). Participants with a healthier diet over the preceding decades had a slower Pace of Aging at baseline and had a lower risk of dementia (**Figure 2, panel A**). Each 1-SD increase in DGAI score was associated with a 0.19 SD (95% CI, -0.24; -0.14) lower DunedinPACE measure, and a HR_C_ of 0.72 (0.58; 0.89) for dementia risk (**Figure 3, panel A**). The association of DGAI score with risk of dementia was attenuated when DunedinPACE was added to the model (HR_C1_ = 0.76 [0.63; 0.91]). Mediation analysis showed that 15% of the effect of DGAI score on the risk of dementia was mediated by DunedinPACE (p = 0.003 for the proportion mediated) (**Figure 3, panel A**).

**Figure 2.**
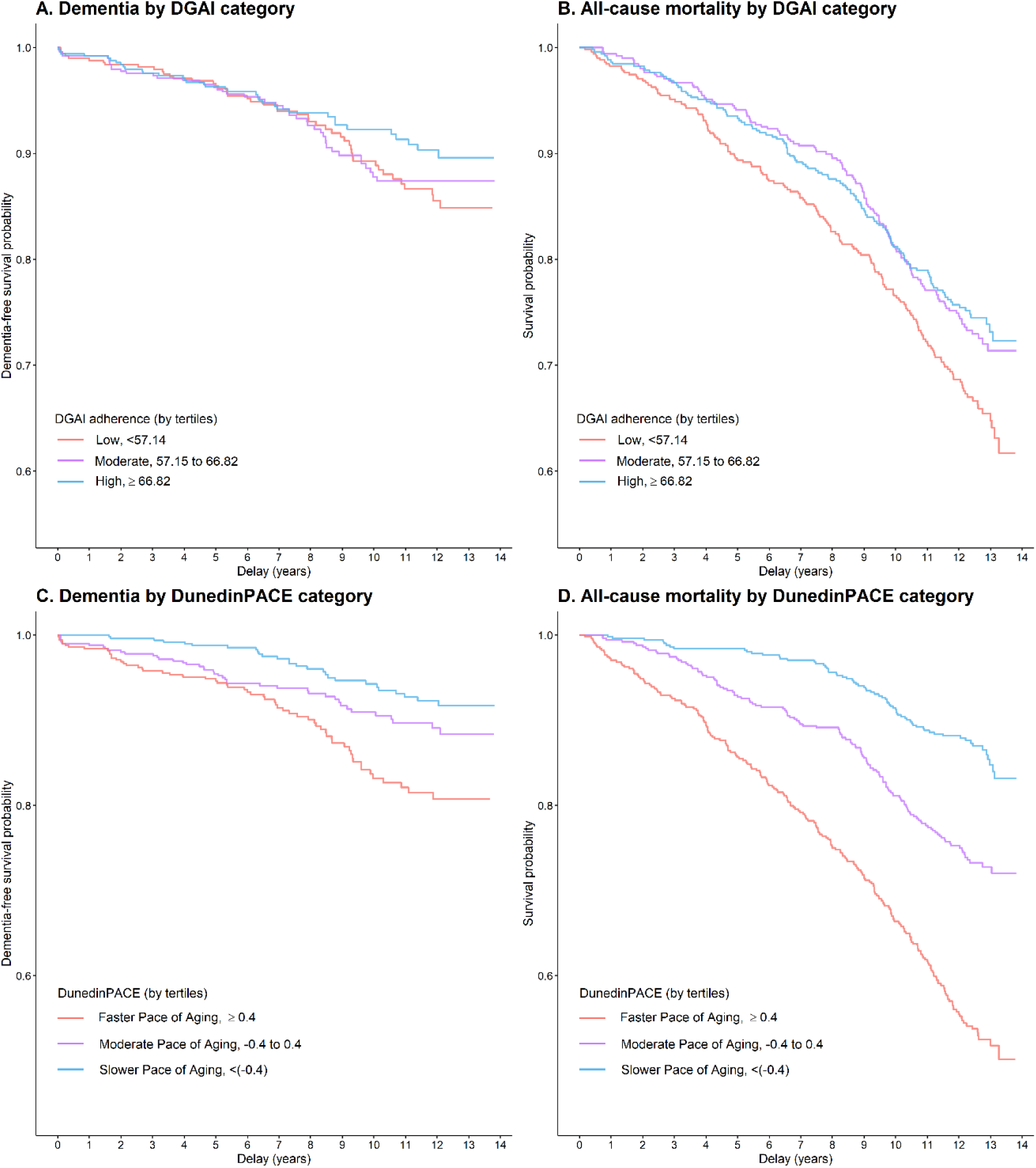
Dementia-free survival and all-cause mortality survival according to categories of DGAI score and of DunedinPACE, estimated by Kaplan-Meier estimator, The Framingham Offspring cohort, 1991-2018 (n = 1,525) Abbreviations: DGAI, dietary guidelines adherence index Kaplan-Meier survival curves for dementia (panels A and C) and for all-cause mortality (panels B and D) displayed by tertiles categories of DGAI score (panels A and B) and DunedinPACE (panels C and D), with delay since baseline as a time scale. Participants with the healthier diet and slowest Pace of Aging (indicated in blue) show lower rates of incident dementia and mortality compared to participants with less healthy diet and faster Pace of Aging (indicated in red).

**Figure 3.**
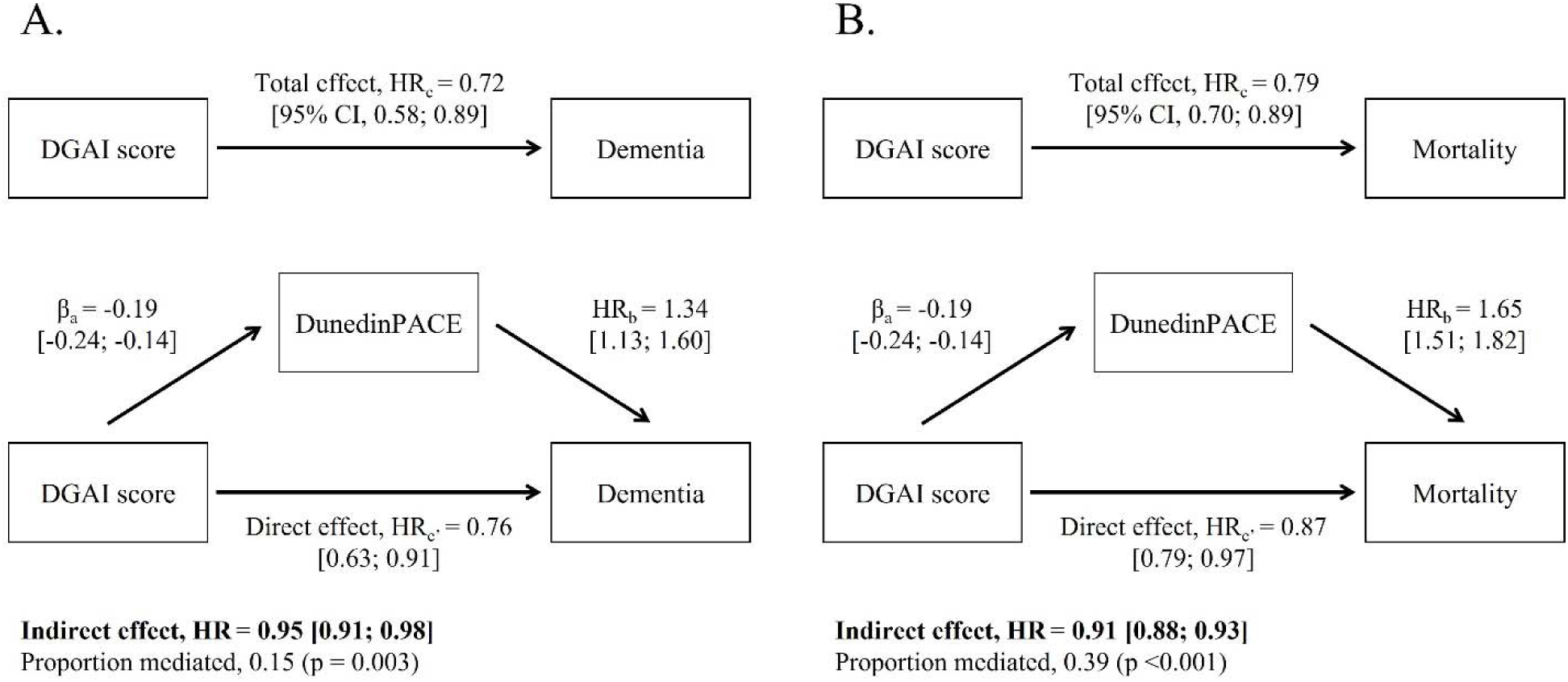
Mediation effect of pace of biological aging on the association of diet with dementia (panel A) and mortality (panel B), The Framingham Offspring cohort, 1991-2018 (n = 1,525) Abbreviations: CI, confidence interval; DGAI, dietary guidelines adherence index; HR, hazard ratio. Mediation analyses were used to examine the indirect effects of DGAI score on dementia or mortality, through DunedinPACE. All models were adjusted for age, age squared, sex, total energy intake, indicator of DNA methylation processing facility. β_a_ indicates the association of DGAI score (continuous, for 1 SD increase) with DunedinPACE, estimated by adjusted linear regression. HR_b_ indicate the associations of DunedinPACE with dementia or mortality, estimated by adjusted Cox proportional hazard models. HR_c_ indicate the associations of DGAI score with dementia or mortality, estimated by adjusted Cox proportional hazard models. The HR_c’_ are the remaining direct effects of DGAI score on dementia or mortality, after controlling for the mediator DunedinPACE in adjusted Cox proportional hazard models. The 95% CIs for indirect effects of DGAI score on dementia or mortality via DunedinPACE were estimated from 10,000 bootstraps sample.

### 3.2. Diet, Pace of Aging, and risk of all-cause mortality

A total of 432 participants (28.3%) died during the follow-up (median, 12.2 years; maximum, 13.6 years). Participants with a faster DunedinPACE at baseline were at increased risk of all-cause mortality over follow-up (**Figure 2, panel D** and **Figure 3, panel B**). Healthier diet was associated with lower risk of mortality, with risk decreased by 21% for each 1-SD increase in DGAI score (HR = 0.79 [0.70; 0.89]) (**Figure 2, panel B** and **Figure 3, panel B**). The association of DGAI score with mortality risk was attenuated when DunedinPACE was added to the model (HR_C1_ = 0.87 [0.79; 0.97]). Mediation analysis showed that 39% of the effect of DGAI score on mortality risk was mediated by DunedinPACE (p <0.001 for the proportion mediated) (**Figure 3, panel B**).

### 3.3 Sensitivity analyses

We conducted sensitivity analysis. Models including covariates for participants’ socio-economic characteristics, *APOE*LJ4 status, lifestyle factors, diabetes and cardiovascular diseases, and DNAm estimates of blood cell composition did not meaningfully change results. Results from covariate-adjusted models are reported in **Supplementary Tables 1 and 2**.

We found no significant modification of associations by sex or *APOE*LJ4 status (all p-values for interactions >0.08). However, as previously reported,^19^ the association of diet with biological aging was moderated by smoking status (interaction p-value = 0.01), such as the association was stronger among ever-smokers (β = -0.20 [-0.28; -0.13]) than never-smokers (β = -0.09 [-0.16; -0.03]) (**Supplementary Figure 1**). We explored whether this interaction might reflect correlation of participants’ smoking histories with their dietary behavior. We repeated our analysis among the subset of participants who had ever smoked, including a covariate for the number of visits at which they self-reported as being a current smoker. DGAI score was negatively associated with smoking history (<0.001) among smokers, and the smoking history-adjusted effect-size estimate of diet on biological aging in smokers (β=-0.12 [-0.19; -0.05]) was similar to the effect-size for non-smokers (**Supplementary Figure 1**).

In addition to long-term dietary habits, we considered healthy diet at each visit to explore dietary exposure at different periods in life, from mid-life (Visit 5, mean age 56.1 ±6.9) to older age (Visit 8, mean age 69.7 ±6.8). The associations of diet with pace of biological aging, dementia and mortality, and the proportions mediated for dementia and mortality were significant and of the similar effect sizes for DGAI scores estimated at all visits (**Supplementary Figure 2**).

As a final test of the robustness of findings, we repeated our analysis using an alternative measure of healthy eating, the Mediterranean Diet Score. This diet score was correlated with the DGAI score (r = 0.63, p <0.001) and yield similar results, with DunedinPACE mediating 15% and 46% of Mediterranean Diet associations with dementia and mortality, respectively (**Supplementary Figure 3**).

There are many proposed methods to quantify biological aging. We focused on DunedinPACE because of prior evidence establishing associations with brain aging and risk of dementia. For comparison purposes, we also report results for two other DNAm measures of aging with robust evidence of association with morbidity and mortality, but mixed evidence of association with brain aging and dementia, the PhenoAge and GrimAge epigenetic clocks. Briefly, patterns of associations were similar for these two clocks, although results for dementia were less consistent and effects were of smaller magnitude relative to DunedinPACE (**Supplementary Table 3**).

## 4. Discussion

Analyzing data from the large population-based Framingham Offspring Cohort, we evaluated the mediating role of biological aging in the relationship of diet with dementia. We found that participants with greater adherence to Dietary Guideline for Americans had a slower pace of biological aging, as measured by the DunedinPACE epigenetic clock, and lower risks of dementia and of all-cause mortality. In turn, faster DunedinPACE was associated with greater risks of dementia and mortality. Mediation analysis indicated that 15% of the healthy-diet’s association with dementia risk was mediated by slower biological aging. The parallel mediation proportion for all-cause mortality was 39%. Associations were robust to potential confounders, including demographic, socioeconomic and lifestyle factors, including smoking status, as well as alternative dietary score or biological aging clocks. The stronger effect of diet on pace of aging observed among smokers may reflect the negative correlation of smoking history with healthy diet and the important contribution of smoking to biological aging.^48, 49^

DNA methylation clocks that aim to measure processes of biological aging have emerged as novel measures of risk for aging-related diseases, including dementia.^22–24^ Initial data suggest that they may provide useful biomarkers at the interface between the environment and aging biology.^50^ However, little is known about how these measures relate to cognitive aging and dementia or the extent to which they are shaped by environmental exposures, such as diet. Our findings, showing associations of the DunedinPACE epigenetic clock with dietary exposure and dementia, suggest that this DNAm measure can provide a tool to better understand how environmental factors, such as diet, may contribute to brain aging.

Healthy diet is associated with preservation of cognitive function and brain integrity with aging.^1–3^ Our findings complement those of other studies in elucidating the role of systemic biological aging in mediating this path. However, DunedinPACE mediated less than half as much of the diet-dementia association as compared to the diet-mortality association. The pathways from diet to dementia are diverse and likely include both direct effects of micro-and macro-nutrients on brain health and indirect effects mediated through metabolic, immune, and cardiovascular health.^51^ The systemic aging processes captured by DunedinPACE may relate primarily to the indirect pathways. The large unexplained fraction of the diet-dementia association may therefore reflect, in part, direct effects of nutrients on brain aging. For example, the brain has the higher metabolic demand relative to other organs consuming ∼20% of glucose-derived energy provided by diet, while accounting for only ∼2% of the body weight.^52^ Nutrients play critical roles in neurotransmission, synaptic functioning, and adult neurogenesis; e.g. omega-3 fatty acids are major components if neuronal membranes.^51, 53, 54^ Some nutrients, such as polyphenols or vitamin A, also have anti-amyloid and anti-tau properties.^51, 55–57^ While there are associations of nutrients with brain structure and white matter integrity phenotypes,^40, 55, 58, 59^ these features of brain aging may also have connections with indirect pathways. In midlife adults, slower Pace of Aging is associated with greater brain volumes and brain vascular health related to dementia risk.^25, 60^ Further research is needed to distinguish direct and indirect effects of diet on brain aging.

We acknowledge limitations. Self-reported dietary data are subject to recall bias, which might cause misclassification. We measured diet from 2-4 validated FFQs over 10 years of follow-up. The use of repeated measures may limit measurement error. Moreover, the beneficial effects of diet on biological aging and dementia risk were consistent for long-term healthy diet, as well as for healthy diet defined within the specific life-course periods of midlife and later life. These observations may reflect overall stable dietary habits across adulthood in healthy individuals,^61^ and suggest that even a single dietary assessment can be useful to evaluate the association of diet with age-related outcomes later in life. There is no gold standard measure of biological aging. DunedinPACE was developed from analysis that integrates rates of age-related decline across multiple organ systems, including metabolic, immune, cardiovascular, pulmonary, renal, hepatic, and periodontal systems.^17^ Nutrition is involved in the regulation of many of these processes. Previous research has shown the involvement of brain metabolism,^5, 62^ inflammation and oxidative processes,^63^ and periodontitis^64^ in the relationship of diet with cognitive aging. Our findings of the mediation role of DunedinPACE on the effect of DGAI score on the risk of dementia hence are consistent with these previous findings, but in a more comprehensive way by capturing multiple systems through DunedinPACE. However, our data cannot distinguish a process in which diet improves organ health, which in turn results in a slower Pace of Aging, from one in which diet promotes healthy aging at the cellular level, and thereby preserves the health and functioning of organs. The Framingham Offspring cohort enrolled mainly White participants. This lack of diversity limits the generalizability of our results. Replication of findings in more diverse cohorts is a priority. As in any observational study, residual confounding may persist despite adjustment for multiple potential confounders in the analysis.

Our study shows that the association of healthier diet with lower risk of dementia is partly mediated by a slower pace of biological aging. If our observations are confirmed in more diverse populations, monitoring biological aging may inform dementia prevention. However, the Pace of Aging pathway does not fully explain the association of diet and dementia, suggesting the presence of other, more direct, pathways. Observational studies well-designed to conduct mediation analysis are needed to investigate direct associations of nutrients with brain aging that may operate independently of systemic biological aging.

## Data Availability

All data produced in the present work are contained in the manuscript.

https://www.ncbi.nlm.nih.gov/projects/gap/cgi-bin/study.cgi?study_id=phs000007.v33.p14

https://www.ncbi.nlm.nih.gov/projects/gap/cgi-bin/study.cgi?study_id=phs000724.v10.p14

