## Supplementary Material for "Diet, pace of biological aging, and risk of dementia in the Framingham Heart Study"

### 1. Dietary assessment

*Adherence to a Mediterranean Diet (MeDi)*

The MeDi score, reflecting adherence to the traditional Mediterranean diet, was calculated from nine food components, with sex-specific medians of energy-adjusted intakes used as cut-off values.[1] For beneficial components (vegetables, fruits, legumes, cereals, fish, and ratio of mono-unsaturated to saturated fats), one point was given for an intake equal or greater than the median. For components presumed to be detrimental (dairy products, and meat), one point was awarded for an intake less than the median. For alcohol, one point was given for a mild-to-moderate consumption (i.e., ]0-1] drink per day for women, and ]0-2] drink per day for men). The total MeDi score ranges from 0 to 9, with higher score indicating greater adherence.
Leveraging the repeated FFQs at each visit, MeDi scores were averaged across four examination cycles (visits 5 [1991-1995], 6 [1995-1998], 7 [1998-2001], and 8 [2005-2008]): 77% of participants had all four FFQs, 20% had three, and 3% had two.

### 2. DNA methylation

*PhenoAge and GrimAge algorithms*

PhenoAge and GrimAge are age clocks derived from DNA methylation patterns of whole-blood physiological variables as predictor of mortality. PhenoAge was developed based on 9 clinical biomarkers (albumin, creatinine, glucose, C-reactive protein, lymphocyte percent, mean cell volume, red cell distribution width, alkaline phosphatase, white blood cell count) and chronological age.[2] GrimAge was created based on 7 plasma proteins (adrenomedullin, beta-2-macroglobulin, cystatin-C, growth differentiation factor 15, leptin, plasminogen activator inhibitor 1, and tissue Inhibitor Metalloproteinases 1), DNA methylation-based estimator of smoking pack-years, chronological age and sex.[3] PhenoAge and GrimAge were calculated using an online calculator (https://dnamage.genetics.ucla.edu/new). The measures were then residualized for chronological age at the time of DNA assessment to derive biological age advancement. For analysis, PhenoAge advancement and GrimAge advancement values were further standardized, so that positive values indicate an advanced state of biological aging and increased risk of diseases and mortality; and negative values indicate a delayed biological aging.

**Supplementary Table 1. Mediation effect of pace of biological aging on the association of DGAI score with dementia, The Framingham Offspring cohort, 1991-2018 (n = 1,525)**

| | Model 2<br>Socio-economic | Model 3<br>*APOEε4* | Model 4<br>Lifestyle | Model 5<br>Diabetes & CVD | Model 6<br>Fully adjusted | Model 7<br>Blood cell count |
|---|---|---|---|---|---|---|
| N dementia cases /total N | 116/1374 | 120/1387 | 124/1512 | 127/1515 | 106/1238 | 129/1525 |
| Total effect, $HR_c$ [95%$CI$]<br>(DGAI score on dementia) | 0.75 [0.59; 0.95] | 0.72 [0.58; 0.88] | 0.75 [0.59; 0.93] | 0.71 [0.57; 0.88] | 0.75 [0.58; 0.94] | 0.71 [0.57; 0.88] |
| Mediator regression, $\beta_a$ [95%$CI$]<br>(DGAI score on DunedinPACE) | -0.16 [-0.22; -0.11] | -0.19 [-0.24; -0.14] | -0.10 [-0.15; -0.05] | -0.18 [-0.23; -0.13] | -0.08 [-0.13; -0.03] | -0.16 [-0.21; -0.11] |
| Mediator effect, $HR_b$ [95% $CI$]<br>(DunedinPACE on dementia, adjusted for DGAI score) | 1.37 [1.13; 1.65] | 1.34 [1.12; 1.61] | 1.34 [1.12; 1.60] | 1.33 [1.11; 1.59] | 1.37 [1.12; 1.66] | 1.28 [1.07; 1.53] |
| Direct effect, $HR_{c'}$ [95% $CI$]<br>(DGAI score on dementia, adjusted for DunedinPACE) | 0.79 [0.64; 0.96] | 0.76 [0.63; 0.92] | 0.77 [0.63; 0.93] | 0.75 [0.62; 0.91] | 0.77 [0.62; 0.94] | 0.74 [0.61; 0.90] |
| Indirect effect, $HR$ [95% $CI$]<br>(DGAI score on dementia, via DunedinPACE) | 0.95 [0.91; 0.98] | 0.95 [0.91; 0.98] | 0.97 [0.94; 0.99] | 0.95 [0.92; 0.98] | 0.98 [0.95; 0.99] | 0.96 [0.93; 0.99] |
| Proportion mediated (*P-value*) | 0.16 (0.02) | 0.15 (0.005) | 0.09 (0.01) | 0.13 (0.004) | 0.07 (0.02) | 0.10 (0.01) |

Abbreviations: *APOEε4*, ε4 allele of the apolipoprotein E gene; CI, confidence interval; CVD, cardiovascular disease; DGAI, dietary guidelines adherence index; HR, hazard ratio.

Mediation analysis was used to examine the indirect effects of DGAI score on dementia, through DunedinPACE. Models were run on participants without missing data for covariates. All models were adjusted for age, age squared, sex, total energy intake, indicator of DNA methylation processing facility (Model 1). Model 2 was adjusted for covariates of model 1, and education and marital status. Model 3 was adjusted for covariates of model 1, *APOEε4* carrier status. Model 4 was adjusted for covariates of model 1, and physical activity index, smoking status and body mass index. Model 5 was adjusted for covariates of model 1, and history of diabetes and CVD. Model 6 was adjusted for all above covariates. Model 7 was adjusted for covariates of model 1, and blood cell counts (CD4 T cells, CD8 T cells, natural killer cells, B lymphocytes, monocytes, and granulocytes).

$\beta_a$ indicates the association of DGAI score with DunedinPACE, estimated by adjusted linear regression. The $HR_b$ indicate the associations of DunedinPACE with dementia, estimated by adjusted Cox proportional hazard models. The $HR_c$ indicate the associations of DGAI score with dementia, estimated by adjusted Cox proportional hazard models. The $HR_{c'}$ are the remaining direct effects of DGAI score on dementia, after controlling for the mediator DunedinPACE in adjusted Cox proportional hazard models. The 95% CIs for indirect effects of DGAI score on dementia via DunedinPACE were estimated from 10,000 boostraps sample.

**Supplementary Table 2. Mediation effect of pace of biological aging on the association of DGAI score with all-cause mortality, The Framingham Offspring cohort, 1991-2018 (n = 1,525)**

| | Model 2<br>Socio-economic | Model 3<br>*APOEε4* | Model 4<br>Lifestyle | Model 5<br>Diabetes & CVD | Model 6<br>Fully adjusted | Model 7<br>Blood cell count |
|---|---|---|---|---|---|---|
| N deaths /total N | 375/1374 | 390/1387 | 426/1512 | 428/1515 | 337/1238 | 432/1525 |
| Total effect, $HR_c$ [95% $CI$]<br>(DGAI score on mortality) | 0.86 [0.75; 0.98] | 0.79 [0.70; 0.89] | 0.87 [0.76; 0.98] | 0.80 [0.71; 0.90] | 0.91 [0.79; 1.05] | 0.80 [0.71; 0.89] |
| Mediator regression, $\beta_a$ [95% $CI$]<br>(DGAI score on DunedinPACE) | -0.16 [-0.22; -0.11] | -0.19 [-0.24; -0.14] | -0.10 [-0.15; -0.05] | -0.18 [-0.23; -0.13] | -0.08 [-0.13; -0.03] | -0.16 [-0.21; -0.12] |
| Mediator effect, $HR_b$ [95% $CI$]<br>(DunedinPACE on mortality, adjusted for DGAI score) | 1.62 [1.46; 1.79] | 1.65 [1.49; 1.82] | 1.55 [1.40; 1.72] | 1.57 [1.43; 1.74] | 1.44 [1.27; 1.62] | 1.59 [1.44; 1.75] |
| Direct effect, $HR_{c'}$ [95% $CI$]<br>(DGAI score on mortality, adjusted for DunedinPACE) | 0.93 [0.83; 1.04] | 0.87 [0.78; 0.97] | 0.90 [0.81; 1.01] | 0.87 [0.78; 0.97] | 0.94 [0.83; 1.06] | 0.86 [0.77; 0.95] |
| Indirect effect, $HR$ [95% $CI$]<br>(DGAI score on mortality, via DunedinPACE) | 0.92 [0.89; 0.95] | 0.91 [0.88; 0.94] | 0.96 [0.93; 0.98] | 0.92 [0.89; 0.95] | 0.97 [0.95; 0.99] | 0.93 [0.90; 0.95] |
| Proportion mediated (*P-value*) | 0.50 (0.02) | 0.38 (<0.001) | 0.29 (0.02) | 0.34 (<0.001) | 0.31 (0.20) | 0.30 (<0.001) |

Abbreviations: *APOEε4*, *ε4* allele of the apolipoprotein E gene; CI, confidence interval; CVD, cardiovascular disease; DGAI, dietary guidelines adherence index; HR, hazard ratio.

Mediation analysis was used to examine the indirect effects of DGAI score on mortality, through DunedinPACE. Models were run on participants without missing data for covariates. All models were adjusted for age, age squared, sex, total energy intake, indicator of DNA methylation processing facility (Model 1). Model 2 was adjusted for covariates of model 1, and education and marital status. Model 3 was adjusted for covariates of model 1, *APOEε4* carrier status. Model 4 was adjusted for covariates of model 1, and physical activity index, smoking status and body mass index. Model 5 was adjusted for covariates of model 1, and history of diabetes and CVD. Model 6 was adjusted for all above covariates. Model 7 was adjusted for covariates of model 1, and blood cell counts (CD4 T cells, CD8 T cells, natural killer cells, B lymphocytes, monocytes, and granulocytes).

$\beta_a$ indicates the association of DGAI score with DunedinPACE, estimated by adjusted linear regression. The $HR_b$ indicate the associations of DunedinPACE with mortality, estimated by adjusted Cox proportional hazard models. The $HR_c$ indicate the associations of DGAI score with mortality, estimated by adjusted Cox proportional hazard models. The $HR_{c'}$ are the remaining direct effects of DGAI score on mortality, after controlling for the mediator DunedinPACE in adjusted Cox proportional hazard models. The 95% CIs for indirect effects of DGAI score on mortality via DunedinPACE were estimated from 10,000 boostraps sample.

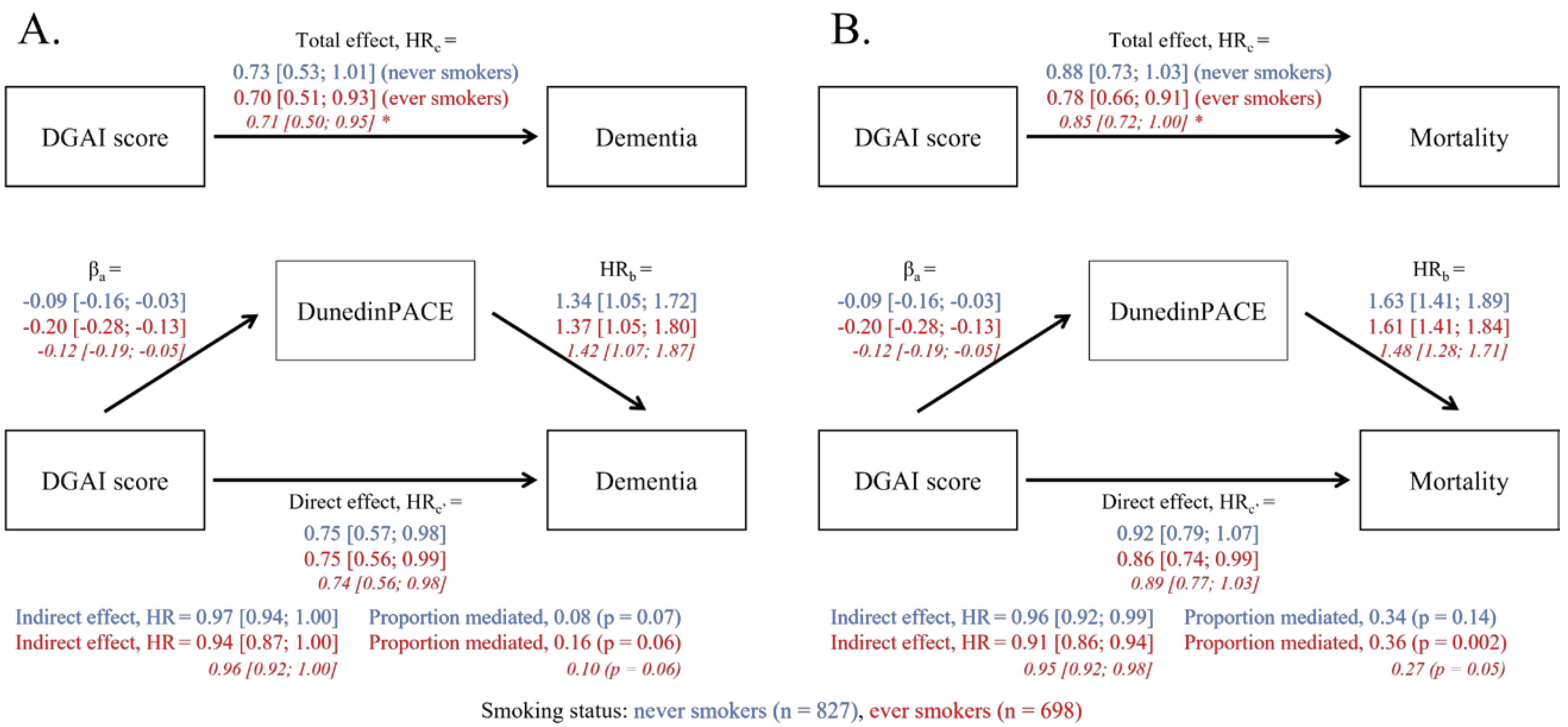


**Supplementary Figure 1. Mediation effect of pace of biological aging on the association of DGAI scores with dementia (panel A) and mortality (panel B), stratified by smoking status, The Framingham Offspring cohort, 1991-2018 (n = 1,525)**

Abbreviations: CI, confidence interval; DGAI, dietary guidelines adherence index; HR, hazard ratio.

Mediation analyses were used to examine the indirect effects of DGAI scores on dementia or mortality through DunedinPACE, separately among never smokers (in blue; n = 827; 73 incident dementia cases and 208 deaths) and ever smokers (in red; n = 698 current or former smokers; 56 incident dementia cases and 224 deaths). All models were adjusted for age, age squared, sex, total energy intake, indicator of DNA methylation processing facility.

* Among ever smokers, a supplementary model was further adjusted for smoking history (computed as the number of visits participants reported being smokers, over the 8 preceding visits from inclusion to study baseline, and weighted for number of visits without missing information for smoking status).

$\beta_a$ indicate the associations of DGAI score with DunedinPACE, estimated by adjusted linear regressions. The $HR_b$ indicate the associations of DunedinPACE with dementia or mortality, estimated by adjusted Cox proportional hazard models. The $HR_c$ indicate the associations of DGAI score with dementia or mortality, estimated by adjusted Cox proportional hazard models. The HRc' are the remaining direct effects of DGAI score on dementia or mortality, after controlling for the mediator DunedinPACE in adjusted Cox proportional hazard models. The 95% CIs for indirect effects of DGAI score on dementia or mortality via DunedinPACE were estimated from 10,000 boostraps sample. A significant interaction was observed between DGAI score and smoking status on the association with DunedinPACE (p = 0.01). No interaction was observed between DGAI score and smoking status or DunedinPACE and smoking status on the associations with dementia and mortality (all p >0.10).

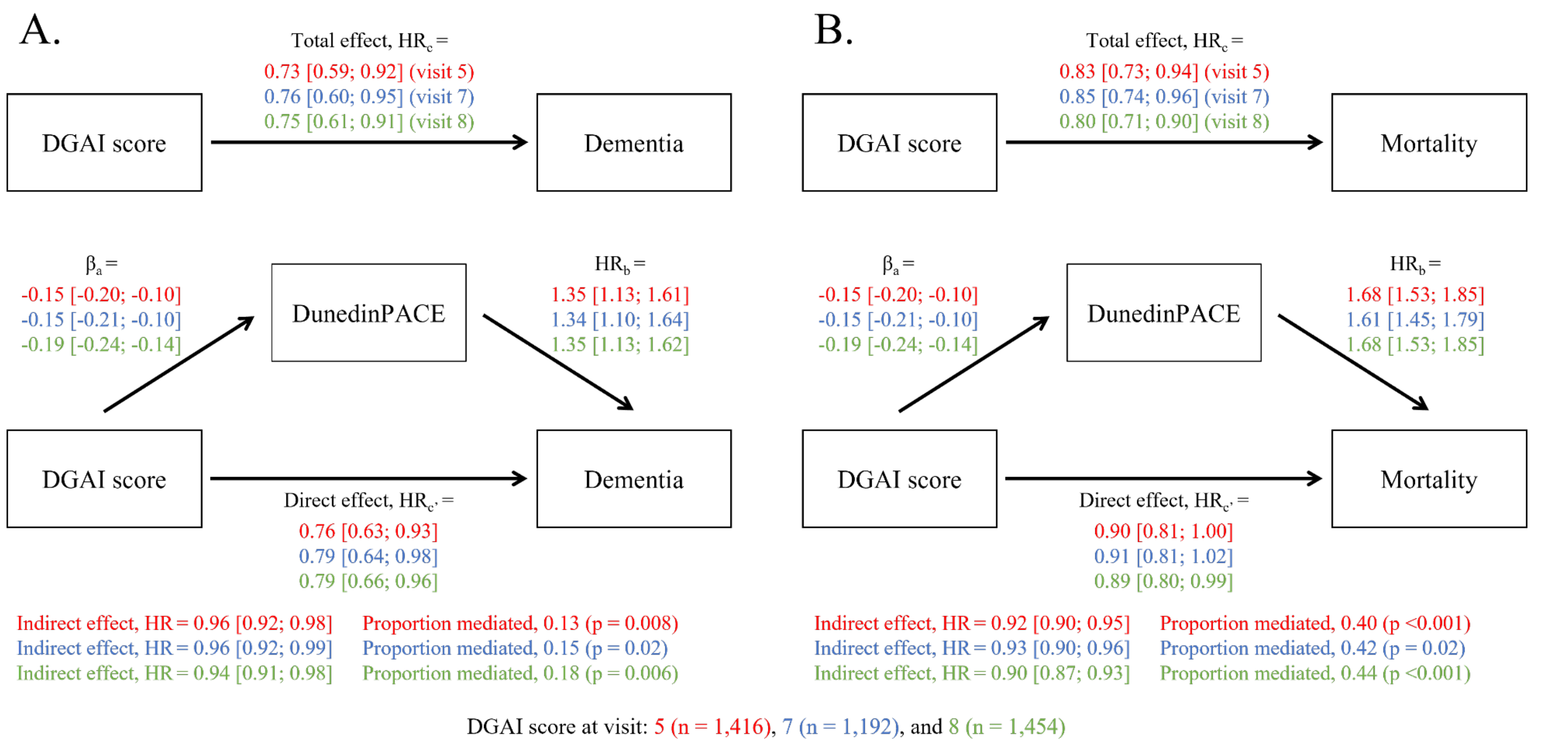


**Supplementary Figure 2. Mediation effect of pace of biological aging on the association of DGAI scores at visits 5, 7, and 8 with dementia (panel A) and mortality (panel B), The Framingham Offspring cohort, 1991-2018 (n = 1,525)**

Abbreviations: CI, confidence interval; DGAI, dietary guidelines adherence index; HR, hazard ratio.
Mediation analyses were used to examine the indirect effects of DGAI scores (computed at visits 5, 7, or 8) on dementia or mortality, through DunedinPACE. The analyses were performed on three sub-samples defined by the availability of DGAI scores at visits 5 (1991-1995; n = 1,416; age range 44-79 years-old; 119 incident dementia cases and 405 deaths), 7 (1998-2001; n = 1,192; age range 51-85 years-old; 101 incident dementia cases and 346 deaths), and 8 (2005-2008; n = 1,454; age range 60-92 years-old; 118 incident dementia cases and 397 deaths). All models were adjusted for age, age squared, sex, total energy intake, indicator of DNA methylation processing facility.
$\beta_a$ indicate the associations of DGAI scores at visit 5, 7, or 8 with DunedinPACE, estimated by adjusted linear regressions. The $HR_b$ indicate the associations of DunedinPACE with dementia or mortality, estimated by adjusted Cox proportional hazard models. The $HR_c$ indicate the associations of DGAI scores with dementia or mortality, estimated by adjusted Cox proportional hazard models. The HRc' are the remaining direct effects of DGAI scores on dementia or mortality, after controlling for the mediator DunedinPACE in adjusted Cox proportional hazard models. The indirect effects of DGAI scores on dementia or mortality via DunedinPACE. 95% CIs were estimated from 10,000 boostraps sample.

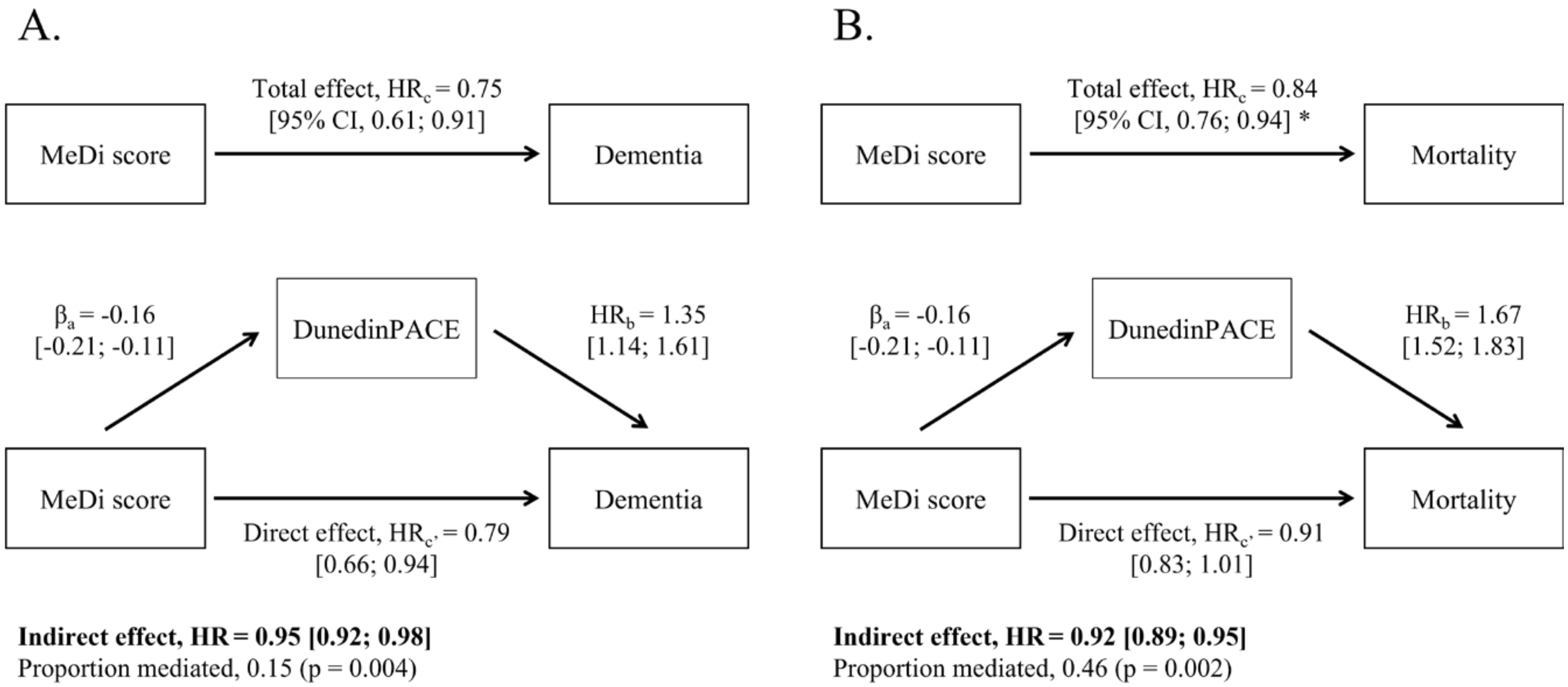


**Supplementary Figure 3. Mediation effect of pace of biological aging on the association of adherence to MeDi with dementia (panel A) and mortality (panel B), The Framingham Offspring cohort, 1991-2018 (n = 1,525)**

Abbreviations: CI, confidence interval; MeDi, Mediterranean diet; HR, hazard ratio.
Mediation analyses were used to examine the indirect effects of MeDi on dementia or mortality, through DunedinPACE. All models were adjusted for age, age squared, sex, total energy intake, indicator of DNA methylation processing facility.
* Adherence to MeDi was considered as a continuous variable (for 1 SD increase); however, the linearity hypothesis for the association of MeDi score with the risk of mortality was not satisfied.
$\beta_a$ indicates the association of MeDi score with DunedinPACE, estimated by adjusted linear regression. The $HR_b$ indicate the associations of DunedinPACE with dementia or mortality, estimated by adjusted Cox proportional hazard models. The $HR_c$ indicate the associations of MeDi score with dementia or mortality, estimated by adjusted Cox proportional hazard models. The $HR_{c'}$ are the remaining direct effects of MeDi score on dementia or mortality, after controlling for the mediator DunedinPACE in adjusted Cox proportional hazard models. The 95% CIs for indirect effects of MeDi score on dementia or mortality via DunedinPACE. were estimated from 10,000 boostraps sample.

**Supplementary Table 3. Mediation effect of alternative biological aging measures on the association of DGAI score with dementia all-cause mortality, The Framingham Offspring cohort, 1991-2018 (n = 1,525)**

| **Biological age** | **PhenoAge** | | **GrimAge** | |
|---|---|---|---|---|
| **Outcome** | **Dementia** | **Mortality** | **Dementia** | **Mortality** |
| Total effect, $HR_c$ [95% $CI$]<br>(DGAI score on outcome) | 0.72 [0.58; 0.90] | 0.80 [0.71; 0.90] | 0.73 [0.59; 0.90] | 0.80 [0.72; 0.90] |
| Mediator regression, $\beta_a$ [95% $CI$]<br>(DGAI score on biological age) | -0.12 [-0.17; -0.07] | -0.12 [-0.17; -0.07] | -0.22 [-0.26; -0.17] | -0.22 [-0.26; -0.17] |
| Mediator effect, $HR_b$ [95% $CI$]<br>(Biological age on outcome, adjusted for DGAI score) | 1.23 [1.04; 1.47] | 1.30 [1.19; 1.43] | 1.13 [0.93; 1.39] | 1.74 [1.59; 1.91] |
| Direct effect, $HR_{c'}$ [95% $CI$]<br>(DGAI score on outcome, adjusted for biological age) | 0.74 [0.62; 0.90] | 0.82 [0.75; 0.91] | 0.75 [0.62; 0.90] | 0.91 [0.82; 1.00] |
| Indirect effect, $HR$ [95% $CI$]<br>(DGAI score on outcome, via biological age) | 0.96 [0.95; 1.00] | 0.97 [0.94; 0.99] | 0.97 [0.93; 1.02] | 0.89 [0.86; 0.91] |
| Proportion mediated (*P-value*) | 0.07 (0.04) | 0.13 (<0.001) | 0.07 (0.26) | 0.52 (<0.001) |

Abbreviations: CI, confidence interval; CVD, cardiovascular disease; DGAI, dietary guidelines adherence index; HR, hazard ratio.

Mediation analysis was used to examine the indirect effects of DGAI score on dementia or mortality, through PhenoAge and GrimAge measures of biological age. All models were adjusted for age, age squared, sex, total energy intake, indicator of DNA methylation processing facility.

$\beta_a$ indicate the associations of DGAI score with PhenoAge and GrimAge, estimated by adjusted linear regressions. The $HR_b$ indicate the associations of PhenoAge or GrimAge with dementia or mortality, estimated by adjusted Cox proportional hazard models. The $HR_c$ indicate the associations of DGAI score with dementia or mortality, estimated by adjusted Cox proportional hazard models. The $HR_{c'}$ are the remaining direct effects of DGAI score on dementia or mortality, after controlling for the mediator PhenoAge or GrimAge in adjusted Cox proportional hazard models. The 95% CIs for indirect effects of DGAI score on dementia or mortality via PhenoAge or GrimAge were estimated from 10,000 boostraps sample.
